# Lung Ultrasound Feature Tracking to Quantify Regional Lung Strain in Mechanically Ventilated Pigs

**DOI:** 10.64898/2026.04.16.26351053

**Authors:** Ruth Walters, Madison Allen, Haley Scheen, Cassidy Beam, Zachary Waldrip, Michaela Singule-Kollisch, Addison Varisco, James Garrett Williams, Daniele De Luca, Brian Michael Varisco

**Affiliations:** University of Arkansas, College of Liberal Arts; University of Arkansas for Medical Sciences, College of Medicine; Arkansas Children’s Research Institute; Division of Pediatric and Neonatal Critical Care, “A.Beclere” Medical Center, APHP-Paris Saclay University, (Paris – France); Pathophysiology and Therapeutic Innovation Unit – INSERM U999, Paris Saclay University (Paris – France)

**Author notes:** These authors contributed equally (Ruth Walters, Madison Allen). **Competing interests** BMV and UAMS Bioventures have applied for a non-provisional patent for the use of lung ultrasound strain measurements for the quantification of lung strain. **Authors’ contributions** RW- 1,2,4,5,6 MA-1,2,4,5,6 MSK-2,3,4,5,6 HS-2,4,5,6 CB-2,4,5,6 ZW-2,4,5,6 AKV-2,4,5,6 DDL-1,4,5,6 BMV-1,2,3,4,5,6. 1-Substantial contributions to the conception or design of the work. 2-Substantial contribution to data acquisition. 3-Substantial contribution to data analysis or interpretation. 4-Drafting the work or reviewing it critically for important intellectual content; AND. 5-Final approval of the version to be published; AND. 6-Agreement to be accountable for all aspects of the work in ensuring that questions related to the accuracy or integrity of any part of the work are appropriately investigated and resolved.

## Abstract

**Background:** In patients requiring respiratory support, clinicians rely on physical exam, radiologic, laboratory, and ventilator-derived measures for the provision of sufficient support while minimizing ventilator and “work of breathing” induced lung injury. Point of care lung ultrasound (LUS) is a widely available tool in hospital and clinic environments. To date, LUS has not been used to evaluate lung strain.

**Methods:** We collected LUS images in four anesthetized, neuromuscularly blocked, and mechanically ventilated pigs being used for another experiment. A feature tracking tool was developed which tracked echo-bright lung structures in ten second clips obtained in triplicate of the right and left, upper and lower lung fields using tidal volumes of 4, 6, 8, 10, and 12 mL/kg. Pleural lines were manually drawn and a program for quantifying lung strain developed with assistance from Anthropic Claude Artificial Intelligence tool. Structures were identified in inspiratory and expiratory frames and tracked bidirectionally with median strain per frame used for calculations.

**Results:** Triplicate measures of lung ultrasound images in four pigs had a median coefficients of variation of 35% (23-47% IQR) and linear modeling of strain with tidal volumes of 4-12 mL/kg showed positive correlation with R^2^ value ranging from 0.89 to 0.97. Strain measurements were similar after bronchial administration of 1.5M hydrochloric acid.

**Conclusions:** Regional lung strain quantification using LUS is a viable and potentially useful tool for respiratory support management.

## Background

Mechanical ventilation is a lifesaving intervention in patients admitted to the intensive care unit with respiratory failure. However, the ventilator can also worsen the severity of lung injury, a phenomenon known as ventilator-induced lung injury (VILI) [1,2]. Strategies to prevent VILI, known as lung-protective ventilation, began in earnest with the publication of the ARDSnet trial, which showed that 6 ml/kg of tidal volume substantially decreased mortality compared with 12 ml/kg in patients with acute respiratory distress syndrome (ARDS) [3]. The concept of VILI evolved further with the finding that the mortality benefit from lower tidal volumes was mediated by a decrease in driving pressure, defined as the difference between plateau pressure and positive end-expiratory pressure (PEEP) [4]. The attempt to encapsulate the energy transferred to the lung by the mechanical ventilator led to the definition of mechanical power (MP) [5]. Mechanical power has since been shown to be positively associated with mortality in mechanically ventilated adults and children with a positive dose-response relationship [6–9]. Driving pressure and mechanical power are critical to our understanding of lung-protective ventilation strategies; however, these strategies rely on ventilator measurements and do not assess the lung structure itself to quantify strain, one of the key pathophysiologic mechanisms of VILI [10]. The ability to quickly assess lung structure and track how that structure changes with each respiratory cycle could provide further insights into how the energy imposed on the lung by the ventilator leads to deleterious structural changes.

Point-of-care ultrasound (POCUS) is ubiquitous and used for numerous applications in the ICU. Lung ultrasound (LUS) has become a crucial bedside tool for diagnosing and monitoring respiratory conditions including pneumothorax, pleural effusion, alveolar-interstitial syndrome, and consolidation. The BLUE protocol established a systematic approach to bedside lung ultrasound in acute respiratory failure [11], and international evidence-based consensus recommendations have systematized acquisition protocols and validated diagnostic accuracy, with sensitivity and specificity for key pathologies comparable to or exceeding chest radiography [12]. Lung ultrasound scoring systems have been developed to semi-quantify lung aeration and monitor the effects of therapeutic interventions [13,14]; global lung ultrasound scores have been validated to independently predict successful liberation from mechanical ventilation in ARDS [15].

Speckle tracking is a well-established echocardiographic computational tool that quantifies myocardial strain [16,17]. Strain is defined as the change in length of a structure divided by its initial length (ε = (L – L_0_) / L_0_). For myocardial strain measurements, the distances between echo-bright structures (features) in the myocardium are tracked frame-to-frame over several cardiac cycles to quantify tissue deformation [18]. Speckle-tracking echocardiography has demonstrated clinical utility in assessing ventricular function across numerous disease states including cardiomyopathies, valvular disease, and cardio-oncology surveillance [19]. The same approach has not been developed for the lung, which presents several additional challenges. Ultrasonography requires that sound waves travel through fluid, and it is the reflection of these sound waves that the transducing probe measures. In the lung, artifacts from air (A-lines) and fluid-filled interlobular septa (B-lines) create horizontal reverberation artifacts or vertical hyperechoic lines respectively [20]. The tubular structures of the lung can move in and out of frame over the respiratory cycle as the lung moves inferiorly during respiration, which is visualized as “lung slide”.

We hypothesize that POCUS of the lung can be used to measure lung strain during mechanical ventilation. We describe the development and initial validation of a lung POCUS method to quantify lung strain using a feature-tracking approach in a porcine model of mechanical ventilation and direct lung injury.

## Methods

### Animal Use and Approval

All experiments were performed in accordance with National Institute of Health (NIH) guidelines in the use of laboratory animals and animal approval was granted by the UAMS IACUC (202400000128).

Ultrasound imaging of pig lungs was done in conjunction with another experiment examining the impact of different mechanical ventilation modes on direct lung injury. In this experiment, four female (39.9 ± 2.6kg) Yorkshire pigs are anesthetized with ketamine bolus and continuous infusion, provided neuromuscular blockade with continuous and intermittent rocuronium, tracheostomized with a 7.0 cuffed endotracheal tube, with vascular access with a 7 French triple lumen central venous catheter and a 3 French carotid artery cannula. Pigs were ventilated in the supine position with Dräger Evita Infinity V500 ventilators for the experiment duration. Pigs were ventilated initially in volume control mode with a PEEP of 5cmH_2_O, respiratory rate of 16 breaths/minute, inspiratory time of 0.8 second. Tidal volumes of 4, 6, 8, 10, and 12 mL/kg were administered with imaging of right and left upper and lower lung regions in triplicate at each volume. A direct lung injury was induced in which 2.5 mL/kg of 1.5 M hydrochloric acid was administered to each of the right and left mainstem bronchi using a bronchoscope, following which injurious mechanical ventilation was induced by exposing the lungs to an end expiratory pressure of 0 cmH_2_O for 10minutes. This was repeated with 1.25 mL/kg of 1.5 M hydrochloric acid administered to each of the right and left mainstem bronchi, followed by 10 minutes of injurious mechanical ventilation. Following injury, ultrasound imaging was repeated.

### Image Acquisition and Preprocessing

Lung ultrasound videos of 8-10 seconds were acquired using a GE BK 5000 with 18L5 TL curvilinear array probe at 7.5 MHz. Exported AVI files were processed as grayscale intensity matrices at 22 frames per second, with each frame extracted sequentially for analysis. The pleural line was manually identified by drawing a line over the pleura over the composite image of 10 random frames. A ten-pixel-tall band centered on the pleural line was designated as the pleural region and excluded from tissue zone analysis. The bottom 5% of the image was cropped to remove data labels, and the remaining tissue depth between the pleural band and cropped boundary was divided into equal thirds, defining Zone 1 (subpleural), Zone 2 (intermediate), and Zone 3 (deep).

### Respiratory Phase Detection

Respiratory phases were identified by modeling mean frame intensity over time. The sinusoidal intensity signal was detrended and windowed using a Hamming function, and a fast Fourier transform (FFT) was applied to identify the dominant respiratory frequency within a physiological range of 10 to 24 breaths per minute (expected: 16). Cardiac oscillations were removed using a 4th-order Butterworth low-pass filter tuned for a heart rate range of 80 to 140 beats per minute. Analysis was restricted to the interval between the first and last completed respiratory cycle.

### Anchor Frame Selection

From detected respiratory extrema, anchor frames were selected at expiratory peaks (high mean intensity) and inspiratory troughs (low mean intensity) based on temporal stability. Frame persistence was quantified as the inverse of pixel-wise variance (i.e. variance in intensity for a given x and y coordinate through all frames) relative to the mean frame, with higher values indicating more stable anatomical positioning. The frames at each respiratory phase transition were designated as anchor frames for centroid initialization and tracking reference points.

### Feature Detection and Centroid Placement

Centroids for feature tracking were identified based on persistence and enforcing minimum spacing between centroids to maximize identification of stable, trackable tissue features. Features were identified in each of three zones using Harris corner detection on gradient images computed with Sobel operators. This approach inherently suppresses responses along linear features such as A-lines and B-lines as it produces high trace but low determinant values. Candidates exceeding the 85th percentile response threshold were selected, with a minimum spacing of 50 pixels enforced to ensure spatial coverage across each zone. Up to 15 candidate centroids were detected per zone.

### Bidirectional Centroid Tracking

Centroids were tracked bidirectionally between consecutive anchor frames using a two-stage cascade method. First, FFT-based cross correlation was performed on a local window around each centroid to estimate coarse displacement. This estimate was then refined using normalized cross-correlation (NCC) template matching with a 25x25 pixel template within a 10-pixel search radius. Frame-to-frame displacement was constrained to 1% of image dimensions to prevent erroneous jumps. Low confidence matches were flagged for imputation. Forward and backward tracks were reconciled by comparing the two independently computed positions. When forward and backward positions agreed within 5 pixels, the average position was used. When they disagreed, the position with higher tracking confidence was selected.

### Motion Model Imputation

Following bidirectional tracking, a consensus motion model was constructed from successfully tracked centroids. This model captured the aggregate displacement pattern across frames, representing the expected tissue motion field. Frames where individual centroids failed tracking (low confidence or flagged positions) were imputed using the motion model, replacing missing positions with model-predicted displacements anchored to each centroid’s last confident position.

### Trajectory Post-Processing

Tracked trajectories underwent spike-based artifact detection using bilateral neighborhood analysis. For each frame, the centroid position was compared against the median position of neighboring frames on both sides (3 frames per side, minimum 2 valid neighbors required). Frames where the displacement from both the left and right neighborhood medians exceeded 5 pixels were classified as tracking artifacts and replaced with linearly interpolated values.

### Quality Assessment and Centroid Selection

Tracking quality was assessed for each centroid based on the proportion of frames with confident (non-imputed) positions. The five highest-quality centroids per zone were selected for strain analysis, with a minimum of three centroids per zone required. Frames from incomplete respiratory cycles at video edges were excluded from all downstream analysis.

### Strain Calculation

Tissue strain was computed from centroid displacement trajectories. For each zone, pairwise Euclidean distances between selected centroids were calculated at each frame. Strain was defined as the fractional change in inter-centroid distance relative to the reference frame: ε = (L - L_0_) / L_0_, where L is the current inter-centroid distance and L_0_ is the reference distance. Zone-level strain was computed as the mean strain across all valid centroid pairs within each zone. Strain values exceeding 0.2 (20%) were considered physiologically implausible since expected lung strain is 1-4% [10] and excluded as tracking artifacts.

### Parameter Tuning

Using minimization of coefficients of variation (CV) as an outcome measure, we varied centroid movement limit, minimum centroid distance, search radius between frames, template size, quality threshold, consistency threshold, and the FFT window to optimize parameters before processing all videos.

### Statistical Analysis

Summary statistics were computed across all processed videos focusing on CV of replicates as a measure of reproducibility and R^2^ of strain vs. tidal volume plots and reproducibility in HCl injured lungs. Correlation of strain values with change in image intensity identified the correlation of change in image intensity (presumably from aerated lung) with strain values. Breath-by-breath comparison of strain within each image was also undertaken as a measure of consistency.

Claude AI (Anthropic, Sonnet 3.5, Opus 4.1 & 4.5 models) was used to aid in computer programming.

### Availability of data and materials

The datasets used and/or analyzed during the current study are available from the corresponding author on reasonable request

## Results

### Conceptual Framework

Before developing the program, we conceptualized image acquisition, processing, and analysis. To assess strain in different lung regions, a linear ultrasound probe is positioned between ribs in these regions to minimize rib shadowing. Tubular lung structures may be imaged in cross-section, obliquely, or longitudinally (Figure 1A). With respiration and movement of the lung with respect to the probe, structures can stay in frame or move out of frame (Figure 1B) which necessitates a method to identify structures that move out of frame and impute location based on other image features. Since the lung moves with each breath, the program must be able to differentiate synchronous movement (no strain) from movement with changes in distance between them (strain, Figure 1C). Images were cropped (Figure 1D) and pleural line drawn in a composite image (Figure 1E) prior to processing.

**Figure 1:**
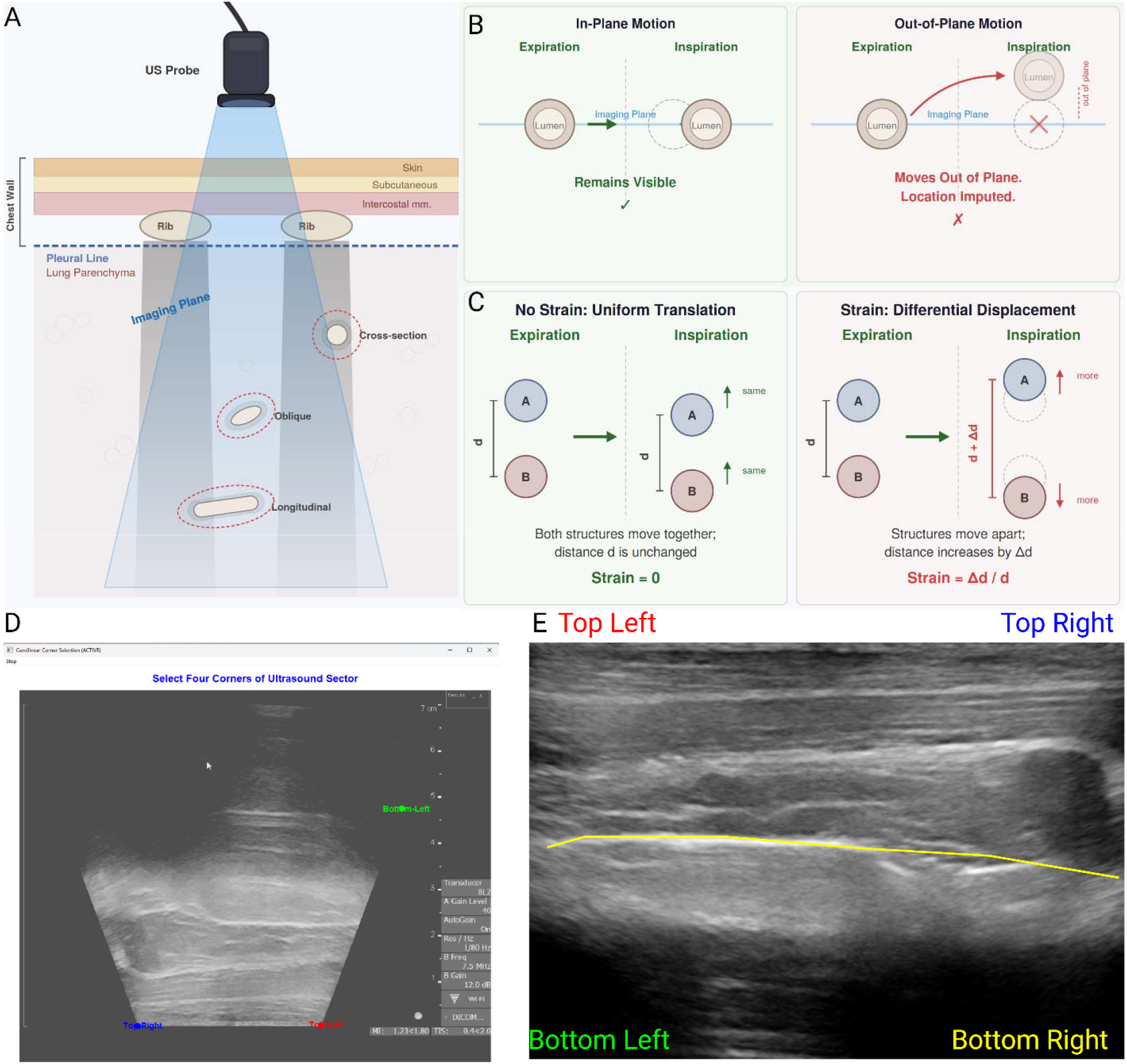
Conceptual Schematic. (A) Lung tissue was imaged by linear ultrasound probe to minimize rib shadows. Lung structures are tubular and could be imaged in cross-section, obliquely, or longitudinally. (B) During respiration, structures can move in-plane and thus be visible throughout the respiratory cycle, or out-of-plane and move in and out of view during the respiratory cycle. In this latter case, the structure is consistently visible during either expiratory or inspiratory phases, but not both. (C) The lung moves during respiration, and movement of lung structures can be due to translation or strain. In translation, structures move uniformly, and in strain, the distance between them changes. In quantifying strain, a distinction must be made between the two. (D) Representative ultrasound image which is inverted. (E) scaled ultrasound image with manually drawn pleural line. Created in BioRender.

### Parameter Tuning

We first needed to determine the proper filter for defining inspiratory and expiratory frames from frame intensity values. We tested first through fourth order Butterworth and Bessel filters in 20 randomly selected videos. While most performed similarly, 2^nd^ order Butterworth filter performed best for videos with clear and less-clear differences between inspiratory and expiratory intensity (Supplemental Figure 1, note that for cardiac oscillations, a 4^th^ order Butterworth Filter was used).

Using 45 sets of triplicate images for tidal volume titration, we performed parameter optimization for centroid movement limit, minimum centroid distance, search radius between frames, template size from which centroid was derived, quality threshold for tracking vs. imputing position, consistency threshold with regards to displacement of the centroid from the starting point, and the FFT window which is the size of the window in which FFT is performed for tracking. We sought to minimize the CV while still yielding strain values from a majority of analyzed videos. Videos in which less than three centroids per zone could be tracked were considered unanalyzable. Since the quality of imaging appeared better in lower lobes and the cardiac motion was greater in left-sided images than right-sided ones, we also evaluated lung regions separately to determine whether there was a region that could not be analyzed due to anatomy. We identified a movement limit of 4%, and a minimum centroid distance of 50 pixels as optimal. The search radius, template size, quality, threshold, and consistency threshold made little difference, and values of 30 pixels, 20 pixels, 0.1, and 0.2 respectively were chosen. The FFT window size had higher variability with larger window sizes and window size of 51 pixels was chosen (Supplemental Figure 2). Lung zone-specific analyses did not yield substantially different parameters. The same optimized parameters were used for analysis of all images.

### Consistency of Strain Measurements

Of the 486 images acquitted in four pigs with different tidal volumes before and after HCl treatment, 426 (87.6%) met the criteria for analysis. All sixty of the excluded videos did not have evaluable strain. There was no clear pattern of exclusion by pig or lung region (Supplemental Figure 3A). As a first assessment of replicability, we evaluated the CV of repeated measures. By pig, median CV ranged from 26.6 to 39.7% (Supplemental Figure 3B). There was no pattern in CV by lung regions (Figure 2A, Supplemental Figure 3C). Smaller tidal volumes tended to have higher CV (Figure 2B). Overall, this data demonstrated acceptable replicability in strain measurements.

**Figure 2:**
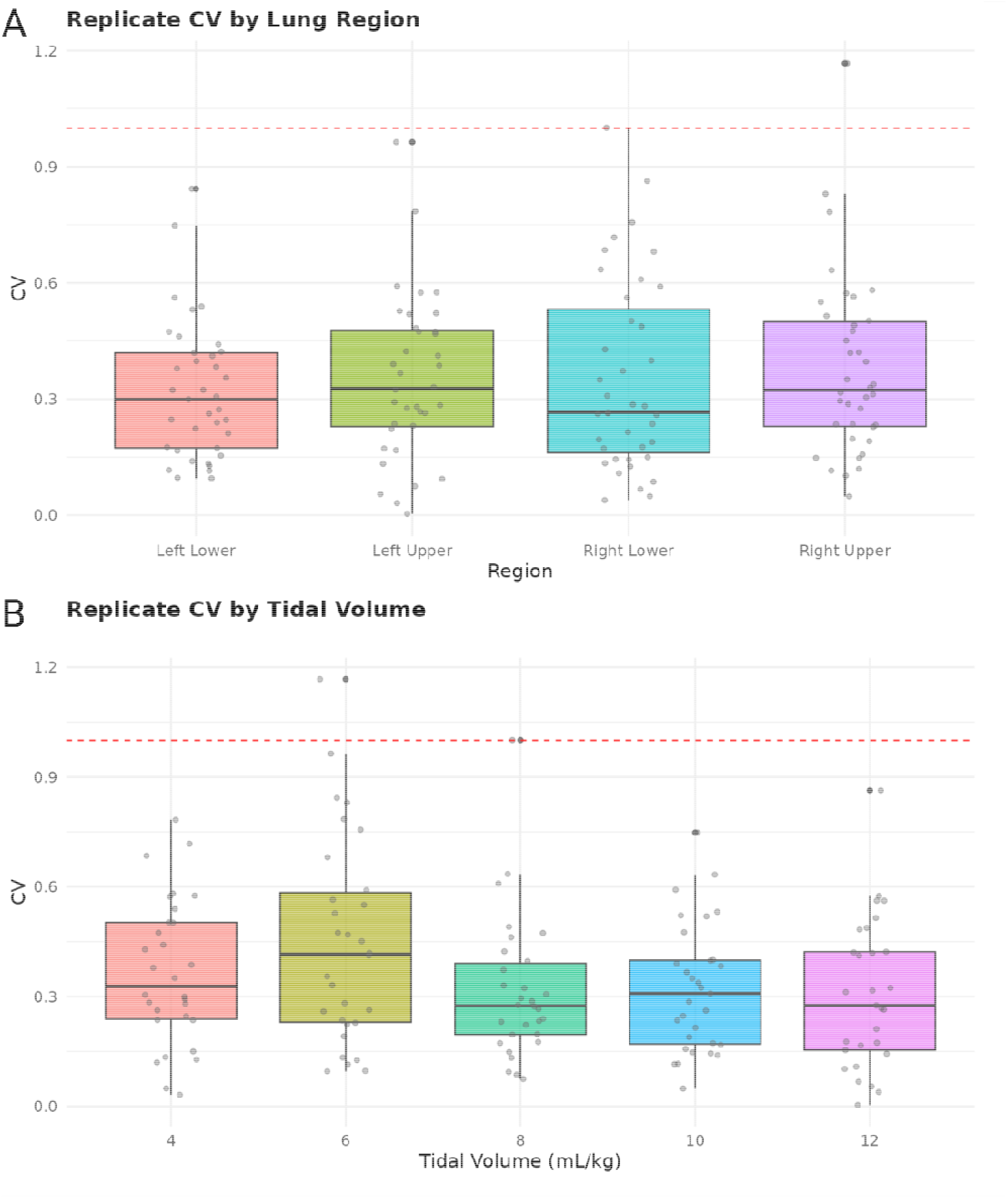
Consistency in Strain Measurement. (A) Coefficients of Variation (CV) were determined from sequential replicate measures of the same animal in the same lung region with the same ventilator settings. CVs did not vary substantially by lung lobe. (B) CV was greater at 6 mL/kg and then 4 mL/kg compared to other tidal volumes. Created in BioRender.

### Validity of Strain Measurements

To establish the within video consistency of strain measurements, we considered both the CV of each breath within each video and the agreement of strain measurements with change in intensity for those breaths. We found a relatively high degree of variation in the breaths within each video (Figure 3A-C). Similarly, when correlating change in image intensity (i.e. level of image “whiteness” or “blackness”) with strain, there were weak but consistently positive correlations for all animals in both baseline and immediately after HCl administration (Figure 3D). These data show consistent correlation but a high level of variability for strain measurements of any individual breath meaning that accurate measurements require repeated measures and multiple breaths per video.

**Figure 3:**
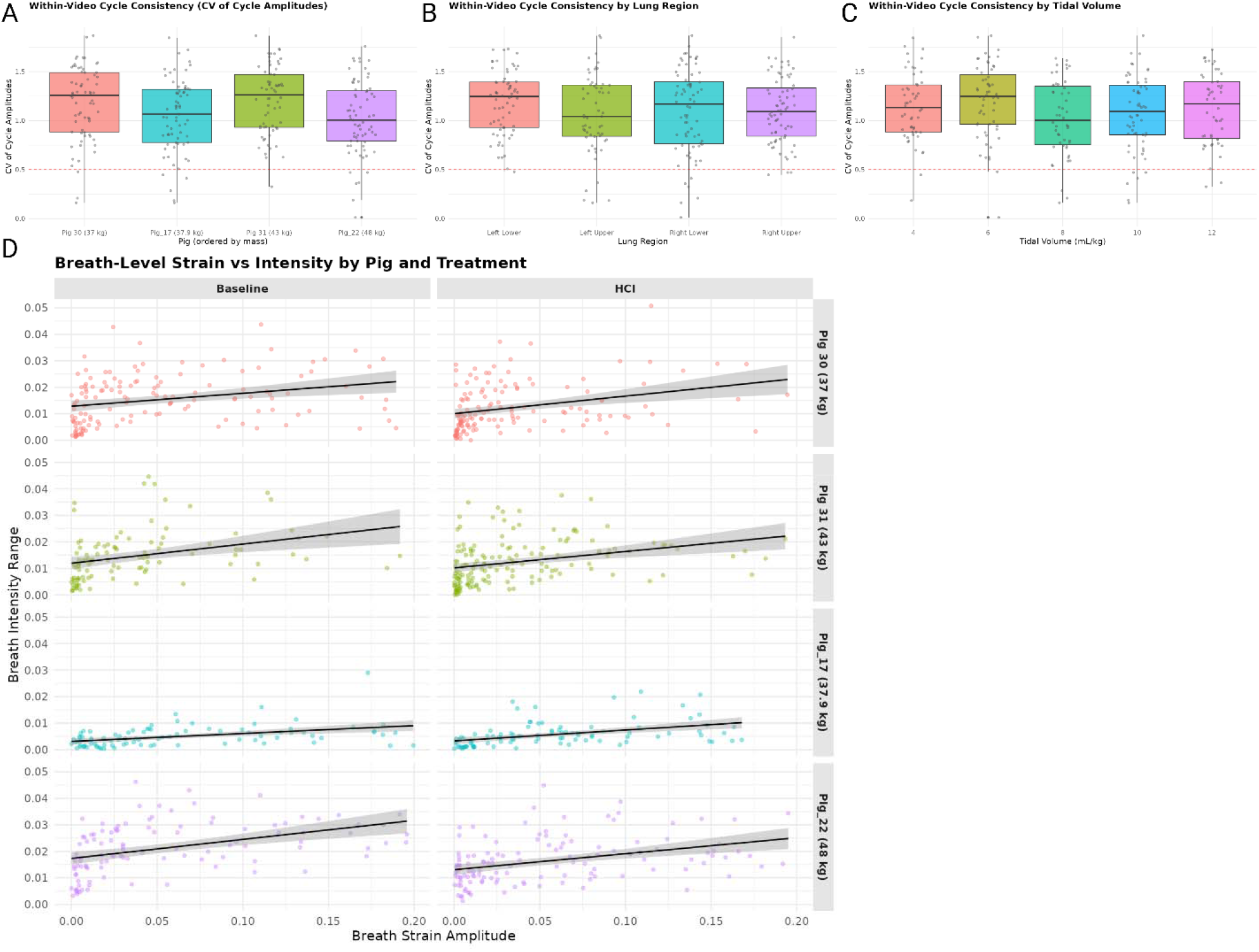
Breath-by-Breath Assessment of Strain. (A) The coefficients of variation for the measured median strain measurement of each breath within a video was calculated and evaluated by animal. There was substantial variability with coefficients of variation around one in analysis by pig (B) by region, and (C) by tidal volume. (D) The change in image intensity for each breath (exhalation intensity – inhalation intensity) was determined for each breath and compared with the median strain for those breaths. A consistent positive but weak correlation was observed for each pig before and after bronchial administration of HCl.

### Tidal Volume and Strain

We used the above parameters to quantify regional lung strain in four sedated, anesthetized, and neuromuscularly blocked pigs, ventilated with 4, 6, 8, 10, and 12 mL/kg in volume control (Figure 4A). Overall and region-specific strain increased with increasing tidal volume (Figure 4B).

**Figure 4:**
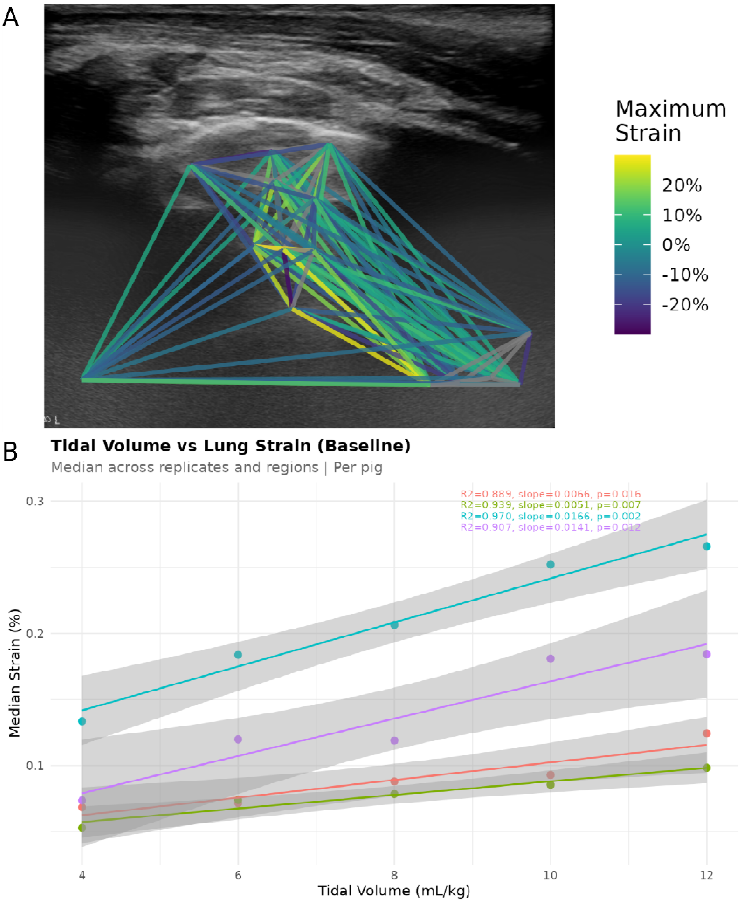
Strain-Tidal Volume Relationships. (A) Representative example of strain calculation. The maximum strain value for each centroid-centroid-pair is shown. Color heatmap scaled lines represent those maximum values with gray lines indicating strain values that were excluded (typically centroid pairs that were close to one another). (B) Four pigs were mechanically ventilated with different tidal volumes in volume control ventilation. All four pigs had increased levels of strain with increasing tidal volume.

### Hydrochloric Acid and Ventilator Mode Studies

The study being used for lung ultrasound imaging involved bronchoscopic administration of 3.75 mL of 1.5 M hydrochloric acid (HCl) to each bronchus via bronchoscope followed by the same titration of tidal volume as described above. Observed strain values were similar before and after HCl administration (Figure 5). These data show that despite variability in individual measurements, reproducible results are achievable with repeated measurements. It is interesting to note that Pig_17 consistently had the highest strain values and least tight correlation to pre-and post-HCl lung strain. This pig also had the most outliers, highest CV (Supplemental Figure 3) and greatest number of videos that required any strain imputation (Supplemental Figure 4).

**Figure 5:**
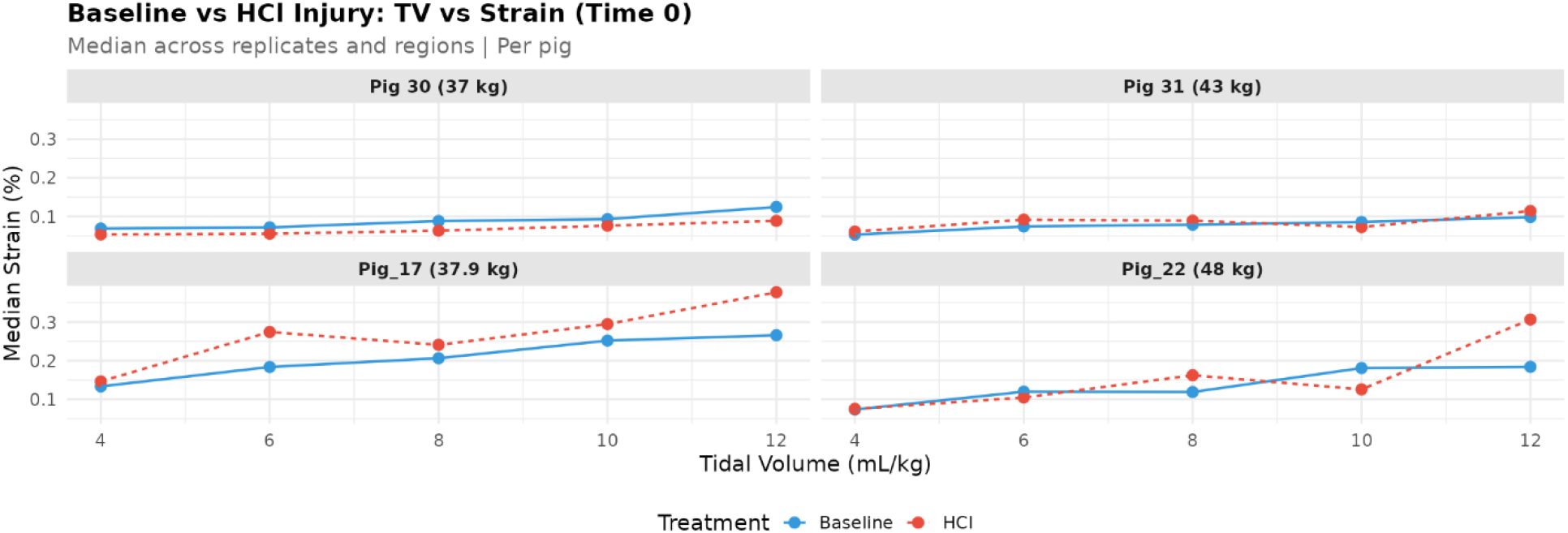
Strain after HCl Injury. Pigs were ventilated with volume control ventilation with the same tidal volumes before (blue circles) and after (red triangles) HCl administration. There was no clear evidence of HCl impacting lung strain immediately after administration.

## Discussion

To the best of our knowledge, this is the first report on the use of LUS to quantify lung strain. We developed our model using sedated, anesthetized, and neuromuscularly blocked pigs with delivery of different tidal volumes and both without and with direct lung injury. We employed several novel techniques that are distinct from the speckle tracking approach used in echocardiography and have thus named the technique feature tracking. These novel approaches include identification of common features in inspiratory and expiratory frames using persistence mapping, ante- and retrograde tracking of features, enforced spacing of features by creation of zones, and imputation of movement in the event of low confidence. This tool could be developed clinically as a stand-alone tool to quantify and intervene upon strain-mediated lung injury in mechanically ventilated and spontaneously breathing patients.

The difference in strain values between pigs is notable. This could be due to technical or pig-specific differences. From a technical perspective animals with greater mass had greater strain values. Imaging sessions were performed on different days, but the same ventilator, technician, ultrasound, and settings were used. It could be that differences between the lungs of each pig accounted for the difference. While the number of alveoli in a pig has not been studied, in healthy adult humans, the number of alveoli ranges from 274-790 million [21]. It may be that measurement differences in lung strain are related to the distribution of tidal volume between fewer or more alveoli. Our finding that pre- and post-HCl injury lungs had similar strain values supports this supposition, but more work, perhaps with stereology, would need to be completed to rigorously test this hypothesis.

This work adds an important dimensional analysis to the emerging field of LUS. LUS can be used as a dynamic imaging technique, that is, it should always be evaluated during patient breathing rather than on still images [22]. Quantitative LUS has also been validated against CT images and several other techniques [23] and reliable guide alveolar recruitment and PEEP titration. Static LUS images cannot provide real-time information about regional lung strain or deformation during the respiratory cycle. Electrical impedance tomography (EIT) represents another bedside regional ventilation monitoring tool, with EIT-derived impedance changes demonstrating strong agreement with CT-measured cyclic lung strain, [28] and EIT-guided PEEP individualization improving oxygenation and reducing alveolar cycling [13]. However, EIT evaluates a single transverse thoracic plane and remains unavailable in most intensive care units globally. Correlation of EIT with LUS lung strain and changes in intensity could be used to develop a dynamic quantification of both aeration and lung tissue movement during respiration.

A recently published editorial advocated for LUS elastography using adapted echocardiography software to assess pleural line strain as a regional deformation surrogate [24], and a simultaneously published open-source algorithm for automated tracking of pleural sliding motion demonstrated technical feasibility for overdistension detection in mechanically ventilated adults [25]. Crucially, however, pleural line tracking measures surface motion rather than intraparenchymal strain. The current approach, by directly tracking echo-bright structures within the lung tissue itself across multiple depth zones, captures deformation that more accurately reflects tissue-level mechanics. Expert commentary has highlighted significant methodological heterogeneity across current feature-tracking platforms, including variation in region-of-interest selection, operator dependence, and proprietary algorithm differences that limit cross-platform generalizability [26]. The development of a purpose-built algorithm for lung parenchymal strain directly addresses this concern.

Several important factors limited our analysis. First, the rib cage anatomy of a pig is different than the human rib cage with wider, more closely spaced ribs. Second, the subcutaneous tissue of a pig thorax is substantially thicker than a human, especially near the neck with the images containing subcutaneous tissue in up to 2/3 of the field, reducing image quality. Third, the ultrasound device available to us is older and newer devices could perhaps provide better image quality. Fourth, due to the open tracheostomy and poorly secured endotracheal tube and open surgical site for internal jugular cannulation, we only imaged anterior lung regions. Lastly, despite our optimization attempts, there was evidence that greater imputation resulted in greater strain values suggesting that the tool could be further optimized.

Future work will need to address several key validation milestones. First, comparison of LUS-assessed parenchymal strain against other physiologic measures —esophageal manometry-derived transpulmonary pressure — will be necessary to establish criterion validity and further evaluate mass-tissue strain findings. Second, the animal-to-human translation challenges identified here (e.g. thicker subcutaneous tissue, wider porcine ribs, limited accessible lung surface) are substantially reduced in humans, particularly in neonates and children where the thin chest wall and smaller intercostal spaces facilitate high-quality parenchymal imaging with high-frequency curvilinear probes. Third, multicenter reproducibility studies will be required to characterize inter- and intra-operator variability [17,18]. Finally, prospective trials will ultimately be needed to determine whether POCUS-guided ventilator adjustments based on regional strain feedback improve clinically meaningful outcomes.

## Conclusion

In summary, we have developed a point of care method for quantifying lung strain using LUS.

## Data Availability

Available upon request

## Acknowledgements

We would like to thank Dr. Douglas Mast at the University of Cincinnati for critiques improving study and manuscript quality.

## List of Abbreviations

VILI: Ventilator Induced Lung Injury
ARDS: Acute Respiratory Distress Syndrome
POCUS: Point of Care Ultrasound
HCl: Hydrochloric Acid
LUS: Lung Ultrasound
PEEP: Positive End Expiratory Pressure
VC: Volume Control
CV: Coefficients of Variation
FFT: Fast Fourtier Transform
NCC: Normalized Cross-Correlation

## References

1. Slutsky AS, Ranieri VM. Ventilator-Induced Lung Injury. New England Journal of Medicine. Massachusetts Medical Society; 2013;369:2126–36. 10.1056/NEJMra1208707

2. Curley GF, Laffey JG, Zhang H, Slutsky AS. Biotrauma and Ventilator-Induced Lung Injury: Clinical Implications. CHEST. Elsevier; 2016;150:1109–17. 10.1016/j.chest.2016.07.019

3. Laffey JG, Kavanagh BP. Ventilation with lower tidal volumes as compared with traditional tidal volumes for acute lung injury. N Engl J Med. 2000;343:812; author reply 813-4. 10.1056/NEJM200009143431113

4. Amato MBP, Meade MO, Slutsky AS, Brochard L, Costa ELV, Schoenfeld DA, et al. Driving Pressure and Survival in the Acute Respiratory Distress Syndrome. New England Journal of Medicine. Massachusetts Medical Society; 2015;372:747–55. 10.1056/NEJMsa1410639

5. Gattinoni L, Tonetti T, Cressoni M, Cadringher P, Herrmann P, Moerer O, et al. Ventilator-related causes of lung injury: the mechanical power. Intensive Care Med. 2016;42:1567–75. 10.1007/s00134-016-4505-2

6. von Düring S, Liu K, Munshi L, Kim SJ, Urner M, Adhikari NKJ, et al. The Association Between Mechanical Power Within the First 24 Hours and ICU Mortality in Mechanically Ventilated Adult Patients With Acute Hypoxemic Respiratory Failure: A Registry-Based Cohort Study. Chest. 2025;168:901–11. 10.1016/j.chest.2025.03.012

7. Bhalla AK, Klein MJ, Modesto I Alapont V, Emeriaud G, Kneyber MCJ, Medina A, et al. Mechanical power in pediatric acute respiratory distress syndrome: a PARDIE study. Crit Care. 2022;26:2. 10.1186/s13054-021-03853-6

8. Wu Y, Liufu R, Wang Y-Y-Q, Chen Y, Li S, Dong R, et al. Association Between Mechanical Power During Prone Positioning and Mortality in Patients With Acute Respiratory Distress Syndrome. Crit Care Med. 2025; 10.1097/CCM.0000000000006811

9. Parhar KKS, Zjadewicz K, Soo A, Sutton A, Zjadewicz M, Doig L, et al. Epidemiology, Mechanical Power, and 3-Year Outcomes in Acute Respiratory Distress Syndrome Patients Using Standardized Screening. An Observational Cohort Study. Ann Am Thorac Soc. 2019;16:1263–72. 10.1513/AnnalsATS.201812-910OC

10. Chiumello D, Carlesso E, Cadringher P, Caironi P, Valenza F, Polli F, et al. Lung Stress and Strain during Mechanical Ventilation for Acute Respiratory Distress Syndrome. Am J Respir Crit Care Med. American Thoracic Society-AJRCCM; 2008;178:346–55. 10.1164/rccm.200710-1589OC

11. Lichtenstein DA, Mezière GA. Relevance of Lung Ultrasound in the Diagnosis of Acute Respiratory Failure*: The BLUE Protocol. CHEST. Elsevier; 2008;134:117–25. 10.1378/chest.07-2800

12. Volpicelli G, Elbarbary M, Blaivas M, Lichtenstein DA, Mathis G, Kirkpatrick AW, et al. International evidence-based recommendations for point-of-care lung ultrasound. Intensive Care Med. 2012;38:577–91. 10.1007/s00134-012-2513-4

13. De Martino L, Yousef N, Ben-Ammar R, Raimondi F, Shankar-Aguilera S, De Luca D. Lung Ultrasound Score Predicts Surfactant Need in Extremely Preterm Neonates. Pediatrics. 2018;142:e20180463. 10.1542/peds.2018-0463

14. Brat R, Yousef N, Klifa R, Reynaud S, Shankar Aguilera S, De Luca D. Lung Ultrasonography Score to Evaluate Oxygenation and Surfactant Need in Neonates Treated With Continuous Positive Airway Pressure. JAMA Pediatrics. 2015;169:e151797. 10.1001/jamapediatrics.2015.1797

15. Haddam M, Zieleskiewicz L, Perbet S, Baldovini A, Guervilly C, Arbelot C, et al. Lung ultrasonography for assessment of oxygenation response to prone position ventilation in ARDS. Intensive Care Med. 2016;42:1546–56. 10.1007/s00134-016-4411-7

16. Voigt J-U, Pedrizzetti G, Lysyansky P, Marwick TH, Houle H, Baumann R, et al. Definitions for a common standard for 2D speckle tracking echocardiography: consensus document of the EACVI/ASE/Industry Task Force to standardize deformation imaging. Eur Heart J Cardiovasc Imaging. 2015;16:1–11. 10.1093/ehjci/jeu184

17. Geyer H, Caracciolo G, Abe H, Wilansky S, Carerj S, Gentile F, et al. Assessment of Myocardial Mechanics Using Speckle Tracking Echocardiography: Fundamentals and Clinical Applications. Journal of the American Society of Echocardiography. Elsevier; 2010;23:351–69. 10.1016/j.echo.2010.02.015

18. Amzulescu MS, De Craene M, Langet H, Pasquet A, Vancraeynest D, Pouleur AC, et al. Myocardial strain imaging: review of general principles, validation, and sources of discrepancies. Eur Heart J Cardiovasc Imaging. 2019;20:605–19. 10.1093/ehjci/jez041

19. Thavendiranathan P, Poulin F, Lim K-D, Plana JC, Woo A, Marwick TH. Use of Myocardial Strain Imaging by Echocardiography for the Early Detection of Cardiotoxicity in Patients During and After Cancer Chemotherapy. JACC. American College of Cardiology Foundation; 2014;63:2751–68. 10.1016/j.jacc.2014.01.073

20. Bhoil R, Ahluwalia A, Chopra R, Surya M, Bhoil S. Signs and Lines in Lung Ultrasonography. Journal of Ultrasonography. 2021;21:225–225. 10.15557/JoU.2021.0036

21. Ochs M, Nyengaard JR, Jung A, Knudsen L, Voigt M, Wahlers T, et al. The number of alveoli in the human lung. Am J Respir Crit Care Med. 2004;169:120–4. 10.1164/rccm.200308-1107OC

22. Raimondi F, Yousef N, Migliaro F, Capasso L, De Luca D. Point-of-care lung ultrasound in neonatology: classification into descriptive and functional applications. Pediatr Res. 2021;90:524–31. 10.1038/s41390-018-0114-9

23. Mongodi S, Cortegiani A, Alonso-Ojembarrena A, Biasucci DG, Bos LDJ, Bouhemad B, et al. ESICM—ESPNIC international expert consensus on quantitative lung ultrasound in intensive care. Intensive Care Med [Internet]. 2025 [cited 2025 May 27]; 10.1007/s00134-025-07932-y

24. Girard M, Garcia D, Carrier FM, Cloutier G. Lung Ultrasound Elastography: New Tools to Personalize Mechanical Ventilation. Am J Respir Crit Care Med. American Thoracic Society - AJRCCM; 2025;211:2214–5. 10.1164/rccm.202502-0406LE

25. Costamagna A, Smit MR, Pivetta E, Persona P, Navalesi P, Pisani L, et al. Tracking pleural sliding motion to assess lung overdistention using an open source algorithm: a proof-of-concept study on lung ultrasound scans. Crit Care. 2026;30:37. 10.1186/s13054-025-05742-8

26. Girard M, Garcia D, Carrier FM, Cloutier G. Lung Ultrasound Elastography: New Tools to Personalize Mechanical Ventilation. Am J Respir Crit Care Med. American Thoracic Society - AJRCCM; 2025;211:2214–5. 10.1164/rccm.202502-0406LE

