## Supplemental Data for "Lung Ultrasound Feature Tracking to Quantify Regional Lung Strain in Mechanically Ventilated Pigs"

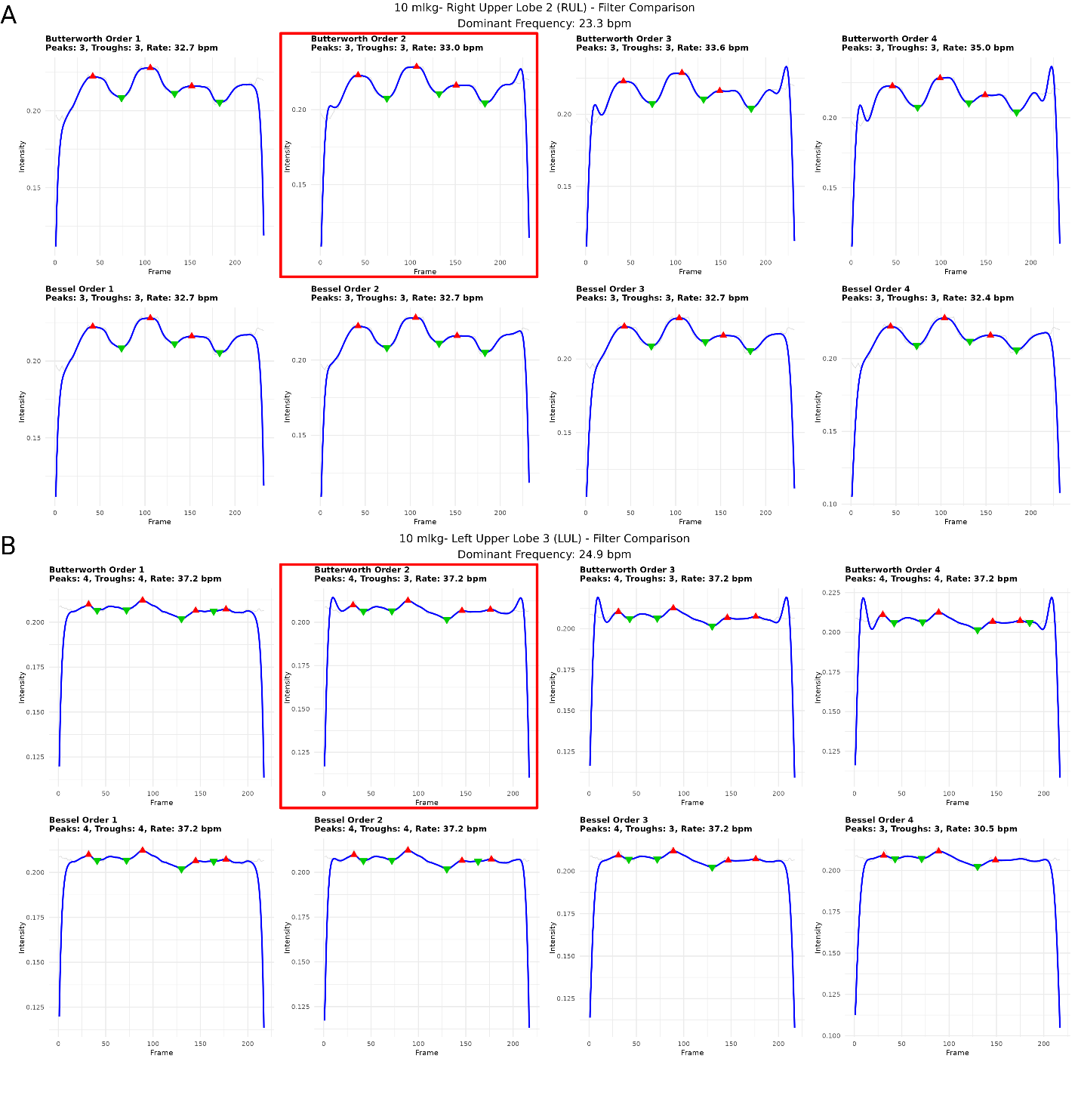

Supplemental Figure 1: Filtering of Intensity Values. (A) Twenty randomly selected videos were selected for testing of different filtering of frame mean intensity values to identify inspiratory (green triangles) and expiratory (red triangles) frames. For images with good differentiation, all tested filters performed similarly. (B) For less clear images, the 2^nd^ order Butterworth (red box) approach had the best discrimination of inspiratory and expiratory valleys and peaks.

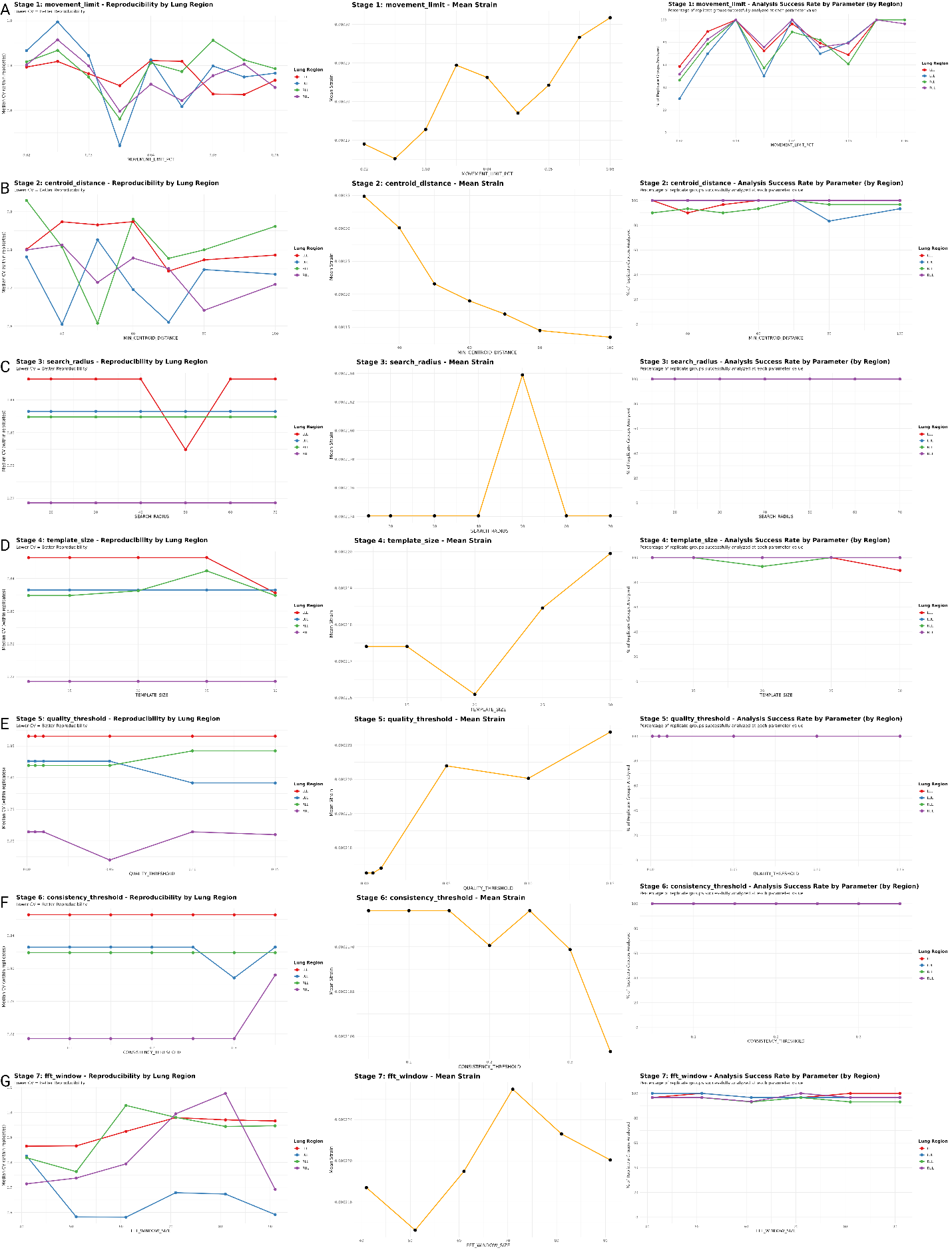

Supplemental Figure 2: Parameter Optimization. (A) Using all images we sequentially titrated acquisition and processing parameters using coefficient of variation (left) fractional strain (middle), and percentage of videos evaluable (right). We evaluated movement limit (B) minimum distance between centroids, (C) search radius between frames, (D) template size, (E) quality threshold, (F) consistency threshold, and (G) fast Fourier transform window size.

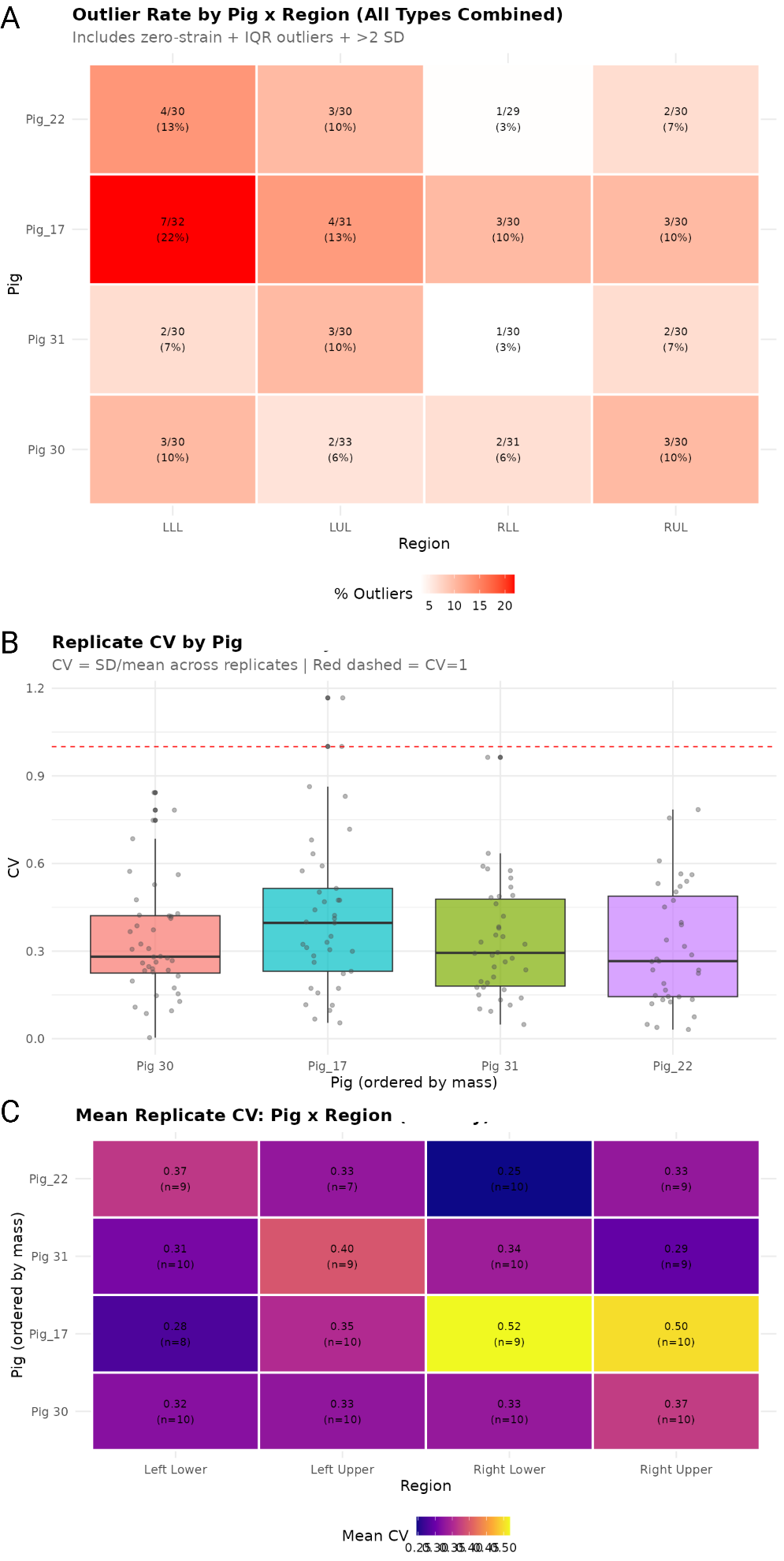

Supplemental Figure 3: Evaluation of Image Strain Consistency. (A) Heatmap showing the relative number of videos that were excluded from analysis because of zero strain or strain more than 1.5 times above the interquartile range. (B) Boxplot of coefficients of variation (CV) for each pig in the study. (C) Heatmap of the CV information by lung region showing that there was no clear pattern in replicability between lung regions.

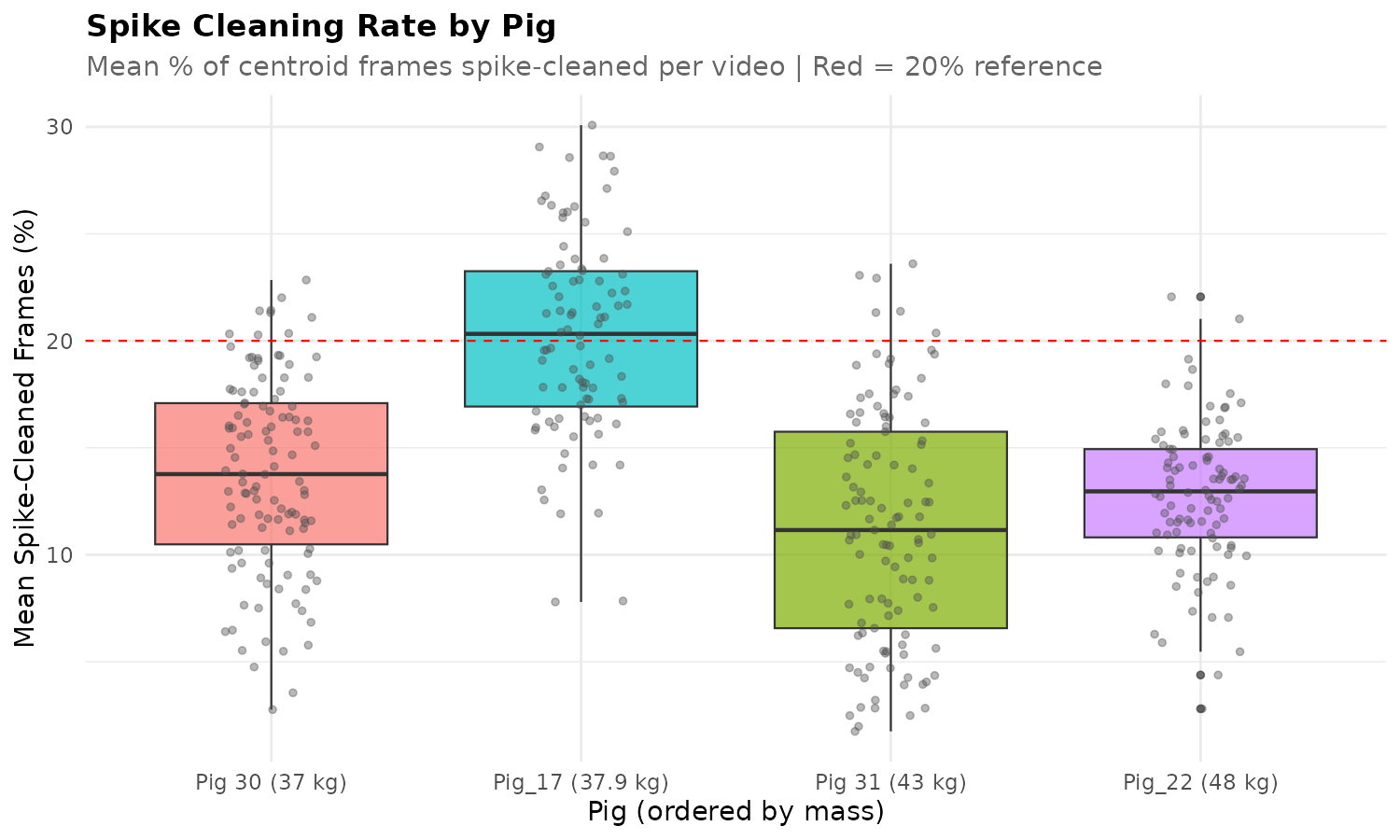

Supplemental Figure 4: Frames with Imputed Strain. Centroid tracking parameters were optimized to prevent jumping, and frames with centroids that exceeded these parameters were marked for imputation based on the movement of other centroids in the video clip. For three of the pigs, about 15% of all videos had at least one frame that required imputation while this number was higher for Pig_17.
